# Prognostic Language and Subsequent Code-Status Limitation After Acute Brain Injury: A Multidatabase Observational Study

**DOI:** 10.64898/2026.08.27.26361534

**Authors:** Alon Gorenshtein, Yosef Adiniaev, Amir Srour, Eyal Klang, Oved Daniel

## Abstract

**Purpose:** Prognostic assessments after acute brain injury are largely narrative, and how prognostic language relates to subsequent care has not been measured at scale. We quantified where it is written and its association with a subsequent code-status limitation.

**Materials and Methods:** Multidatabase observational study of adults with acute brain injury or a related neurologic emergency, using MIMIC-IV (2008-2019; discharge summaries and radiology reports) and a timestamped MIMIC-III cohort (notes and code-status orders). The exposure was documented prognostic language; outcomes were its association with a subsequent full-code-to-limitation transition, note-stream location, and completeness of documented command-following relative to structured Glasgow Coma Scale (GCS) motor scores.

**Results:** Among 31,993 admissions (27,054 patients; median age, 69 years; 54.9% male), prognostic language in the timestamped cohort (MIMIC-III) was associated with a subsequent code-status limitation after multivariable adjustment (adjusted hazard ratio, 4.3; 95% CI, 2.9-6.5; unadjusted 14-day cumulative incidence, 40% vs 8.5%), including the comfort-measures component (3.9), a higher-risk subgroup (4.4), and after acute-physiology adjustment (4.1); the association was concentrated in the first 3 days. Non-prognostic severity language showed no comparable association (hazard ratios, 1.1-1.3). Prognostic language localized almost entirely to the narrative (4.9% of discharge summaries vs 0.015% of radiology reports); command-following was undocumented in 55.7% of summaries, and no final-24-hour GCS motor score was charted in 72.8%.

**Conclusions:** Documented prognostic language after acute brain injury was written in the narrative, not structured fields, and was associated with a subsequent code-status limitation after multivariable adjustment. This observational association cannot establish causation but warrants prospective study.

**Key Points:** *Question:* In routine records of acute brain injury, where is prognostic language written, and is it associated with a subsequent transition to a limited code status?

*Findings:* Across 31,993 admissions, prognostic language was detected in 4.9% of discharge summaries but almost never in radiology reports; in a timestamped cohort it was associated with a subsequent full-code-to-limitation transition after multivariable adjustment (hazard ratio, 4.3; 4.1 with added acute physiology), an association not seen for far more common non-prognostic severity text and concentrated in the first 3 days. This observational association cannot establish causation.

*Meaning:* Prognostic language lives in the treating team’s narrative and is closely coupled with subsequent code-status decisions, warranting prospective study.

## Introduction

Assessments of prognosis and observations of whether a patient responds at the bedside inform decisions to continue or limit life-sustaining treatment after acute brain injury.[1–6] Withdrawal of life-sustaining treatment accounts for most in-hospital deaths after severe traumatic brain injury and cardiac arrest, and concern that an early pessimistic prognosis can shape the intensity of subsequent care is longstanding in neurocritical care.[1–6] A substantial minority of behaviorally unresponsive patients retain covert cognitive function, which sharpens the stakes of any assessment that treats the absence of a bedside response as the absence of retained function.[7–14]

Both judgments share a documentary property that has received little empirical attention: they are recorded in narrative text. A structured record can store a Glasgow Coma Scale (GCS) component, a sedation score, or a code-status value, but it has no field for value-laden prognostic language and incompletely captures the narrative of responsiveness. Prior work has described how clinicians communicate prognosis to families,[15–17] and natural-language-processing studies of radiology have extracted findings and diagnostic certainty,[18,19] but the prevalence and note-stream location of prognostic language itself, and its relationship to documented responsiveness and to subsequent care, have not been measured at scale.

We therefore measured, across adults with acute brain injury or a related acute neurologic emergency, where prognostic language is written (Aim 1), how completely command-following is documented and how narrative and structured records correspond (Aim 2), and how prognostic language and documented responsiveness co-occur (Aim 3), using MIMIC-IV, whose linked text comprises only discharge summaries and radiology reports. Because that corpus is not time-stamped, we then used MIMIC-III, which retains timestamped progress notes and code-status orders, to place prognostic language on the admission timeline and to test whether it was associated with a subsequent transition from full code to a code-status limitation (do-not-resuscitate, do-not-intubate, or comfort measures) after multivariable adjustment (Aim 4; Figure 1). The study is observational; the code-status association is adjusted but cannot establish causation.

**Figure 1.**
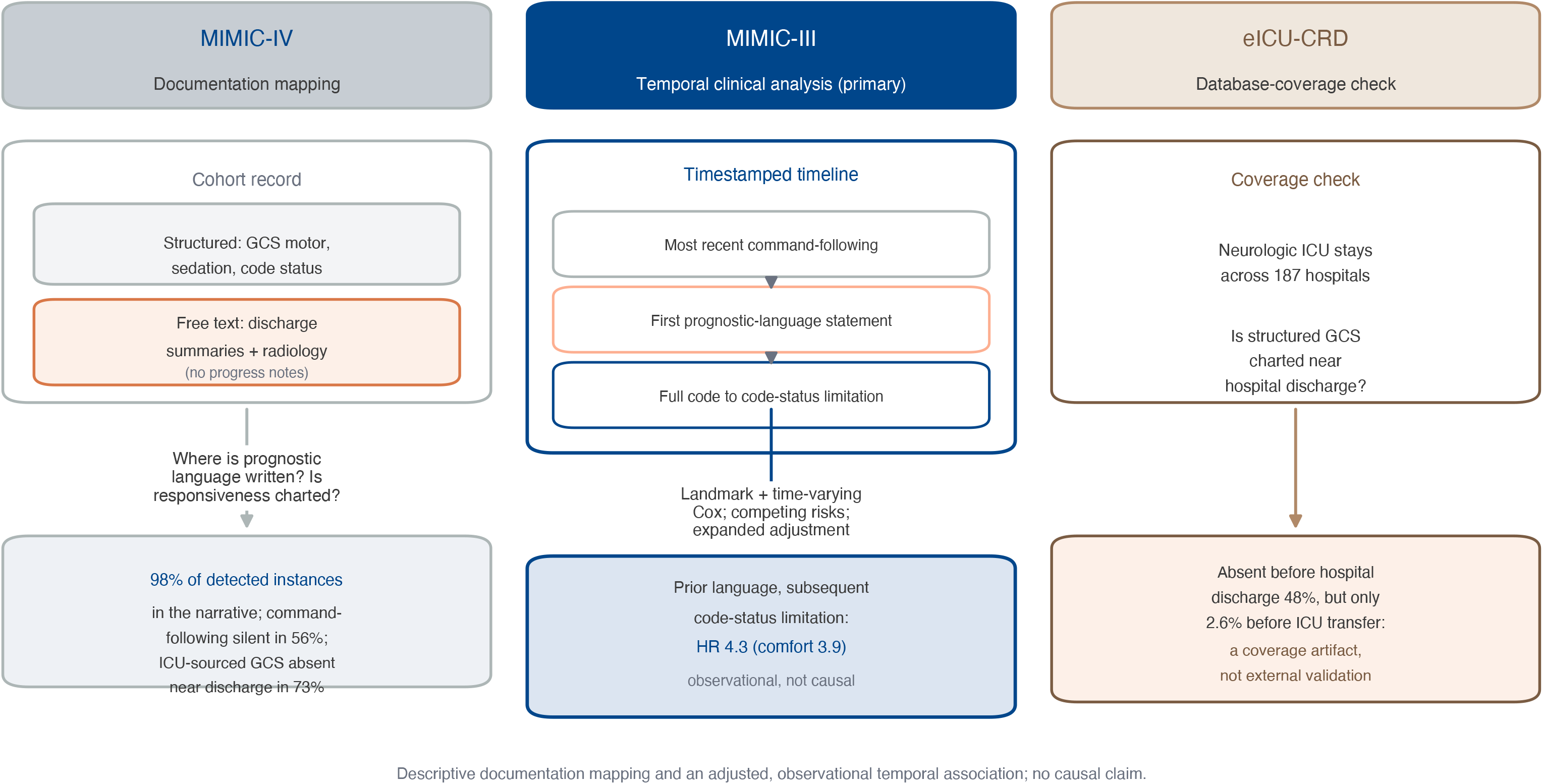
Three-database study design. In MIMIC-IV (documentation mapping), the acute brain injury record comprises a structured layer (Glasgow Coma Scale motor, sedation, and code-status fields) and a free-text layer limited in this corpus to discharge summaries and radiology reports (no progress, consultation, or goals-of-care notes); we measure where prognostic language is written and how completely responsiveness is charted (Aims 1-3). In MIMIC-III (temporal clinical analysis), timestamped notes and code-status orders place the first prognostic statement on the admission timeline and support landmark and time-varying models of subsequent code-status limitation with expanded adjustment (Aim 4). eICU-CRD provides a multicenter check on structured-charting coverage near discharge. The documentation analyses are descriptive; the code-status association is adjusted but observational.

## Methods

### Data source and cohort

We used MIMIC-IV version 3.1, a deidentified critical-care database from a single academic medical center covering 2008 to 2019,[20] linked by admission and patient identifiers to MIMIC-IV-Note version 2.2. The linked corpus contains discharge summaries and radiology reports for these admissions; it does not contain progress notes, consultation notes, or dedicated goals-of-care notes, and all claims are scoped to the two available streams. The study followed the STROBE and RECORD reporting guidelines.[21,22] Analysis of the deidentified, publicly available database was determined not to constitute human-subjects research by the data custodians; the institutional data-use agreement governed access. The analysis code is available at https://github.com/Alon-Gorenshtein/Progsontic-unstructured-.

We included all admissions with an acute brain injury or related acute neurologic emergency identified by International Classification of Diseases, Ninth and Tenth Revision codes for nine phenotypes (anoxic-ischemic injury, cardiac arrest, intracerebral, subarachnoid, and subdural hemorrhage, acute ischemic stroke, traumatic brain injury, status epilepticus, and central nervous system infection; eTable 1). When more than 1 phenotype was coded, a single primary phenotype was assigned by a fixed acuity priority, with co-occurring phenotypes retained as flags (eTable 10); a sensitivity analysis excluded cardiac-arrest admissions without a concomitant anoxic-ischemic diagnosis (eTable 11). Aim 1 used admissions with a discharge summary or head-imaging report; Aim 2, those with both a discharge summary and structured GCS motor charting; Aim 3, those with a discharge summary. A secondary, clinically defined severe-injury subgroup comprised admissions with an intensive care unit (ICU) stay plus mechanical ventilation or a hemorrhagic, anoxic-ischemic, or cardiac-arrest phenotype (11,366 admissions; 28.7% mortality), reported as a sensitivity analysis in the population where prognostic language is not rare.

### Prognostic-language and responsiveness extraction

We defined prognostic language as a value-laden statement about a patient’s expected survival or neurological recovery, distinguished from two contrasting registers that are common in clinical text but are not prognostic-language statements: radiographic severity description (eg, “midline shift,” “large hemorrhage”) and diagnostic-certainty hedging (eg, “consistent with,” “cannot exclude”). A rule-based extractor captured prognostic language in seven groups (eMethods; eTable 3) with negation, family-history, procedural, organ-specific, and irrealis-mood guards, and was frozen before evaluation. Narrative command-following was extracted from discharge summaries with negation-aware logic, classifying each summary as documenting command-following, documenting its explicit absence, both, or neither.

We characterized both extractors against analyst reference labels from stratified random samples, then had two physicians independently adjudicate the same excerpts blinded (eTable 4). Against the two-physician consensus, design-weighted precision, recall, and specificity were 0.91, 0.85, and 0.996 for prognostic language and 0.96, 0.91, and 0.98 for command-following. Because sensitivity and precision are imperfect, reported values are detected prevalences whose net bias direction is unknown.

Documented command-following is a sign of behavioral responsiveness, not of consciousness; we report documented responsiveness throughout and do not infer the underlying level of awareness. Structured command-following was defined as a GCS motor score of 6 (“obeys commands,” item 223901).[23] We compared layers two ways: an unaligned stay-level comparison (any narrative statement vs structured command-following ever recorded during the admission) and a time-matched comparison (the end-of-stay narrative examination vs structured GCS in the final 24 hours before the terminal event). Preserved responsiveness was any of narrative command-following, narrative purposeful responsiveness (visual pursuit or localization), or a structured GCS motor score of 6; “late” responsiveness required the last-24-hour structured measure or the discharge-examination narrative to be positive.

### Statistical analysis

Prevalences are reported with Wilson 95% CIs, and, because patients could contribute more than 1 admission, with patient-clustered bootstrap CIs and a first-admission-only sensitivity analysis (eTable 8). Prognostic-language prevalence across phenotypes was tested with the χ^2^ test and summarized with Cramér V; the phenotype-level relationship between prognostic-language prevalence and mortality used the Spearman correlation. Layer agreement used the Gwet AC1 statistic[24] and Cohen κ, with the McNemar test for directional differences. As a construct-validity check, we tested how far structured features predicted the presence of prognostic language (5-fold cross-validated logistic regression and gradient boosting; area under the receiver operating characteristic curve [AUROC] and Brier score; eTable 7). The Benjamini-Hochberg procedure[25] controlled the false discovery rate across the primary family of tests (2-sided P < .05). Analyses used Python 3.9 (pandas, scikit-learn, SciPy, statsmodels, lifelines).

### Timestamped analysis and code-status limitation (MIMIC-III)

Because MIMIC-IV-Note lacks timestamped progress notes, we repeated the extraction in MIMIC-III version 1.4, a deidentified database from the same institution (2001-2012) that retains timestamped physician, nursing, and consultation notes and code-status orders,[26] applying the identical, frozen extractors; their precision in these note types was confirmed by blinded physician adjudication (eTable 14). We placed the first prognostic-language statement on the admission timeline and defined the outcome as the first structured transition from full code to a limitation (do-not-resuscitate, do-not-intubate, or comfort measures), analyzed as a composite and, because these decisions differ clinically, by component (comfort measures; resuscitation orders without comfort measures). In cause-specific Cox models at 24- and 48-hour landmarks, restricted to admissions alive with an observed full-code state at the landmark (not assumed by default), and in a whole-stay time-varying Cox model in which the exposure became active only at the first statement (avoiding immortal-time bias, with baseline covariates), we estimated the hazard of a subsequent code-status limitation for admissions with versus without prior documented prognostic language, treating death and discharge as competing events. Models adjusted for age, primary phenotype, worst GCS motor score, mechanical ventilation, deepest Richmond Agitation-Sedation Scale value, the van Walraven comorbidity index, metastatic cancer, dementia, chronic liver and renal disease, emergency admission, pre-landmark intensive care unit hours, and database source; a further model added pre-landmark acute physiology (worst serum creatinine, bilirubin, platelets, lactate, and blood urea nitrogen, arterial oxygen tension, and any vasopressor exposure) from the laboratory and infusion tables (a full APACHE score was not computable). Models were unpenalized, with confidence intervals clustered by patient; proportional hazards were tested (Schoenfeld residuals) and, where violated, a piecewise hazard split at 72 hours was fitted. Sensitivity analyses included the physiology-adjusted model, an incidence-density risk-set matched analysis (controls sampled at each statement time with equal covariate windows), a specificity comparison in which the exposure was replaced by non-prognostic text registers (radiographic severity, diagnostic-certainty hedging, and generic critical-illness acuity), a higher-risk neurologic subgroup, covariates measured before the statement, documentation-volume adjustment, strata by documentation era and phenotype, first-admission and 12-hour-interval analyses, and an E-value[27] (eMethods; eTables 15, 18-20).

## Results

### Cohort

The cohort comprised 31,993 admissions from 27,054 patients (median age, 69 years; 54.9% male; in-hospital mortality, 13.0%), most commonly acute ischemic stroke (31.9%) and traumatic brain injury (28.6%). A discharge summary was available for 23,239 admissions (72.6%; the documentation-mapping cohort, characterized in eTable 17), 177,855 radiology reports were linked (41,205 head or brain studies), and 12,007 admissions had both a discharge summary and structured GCS motor charting. The timestamped MIMIC-III cohort comprised 11,079 admissions; its 24-hour landmark subset (n = 5,493) is characterized in Table 1.

**Table 1.**
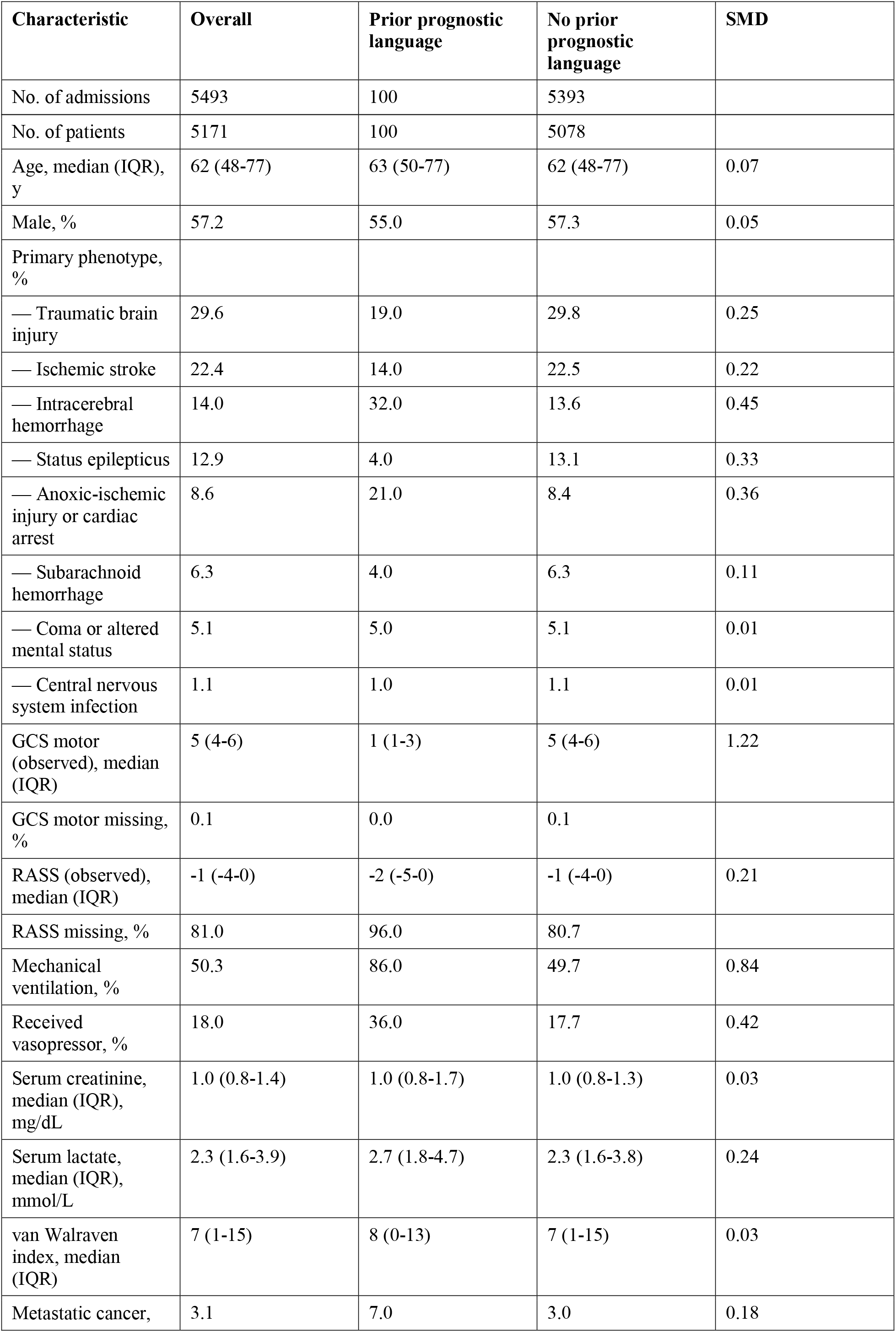
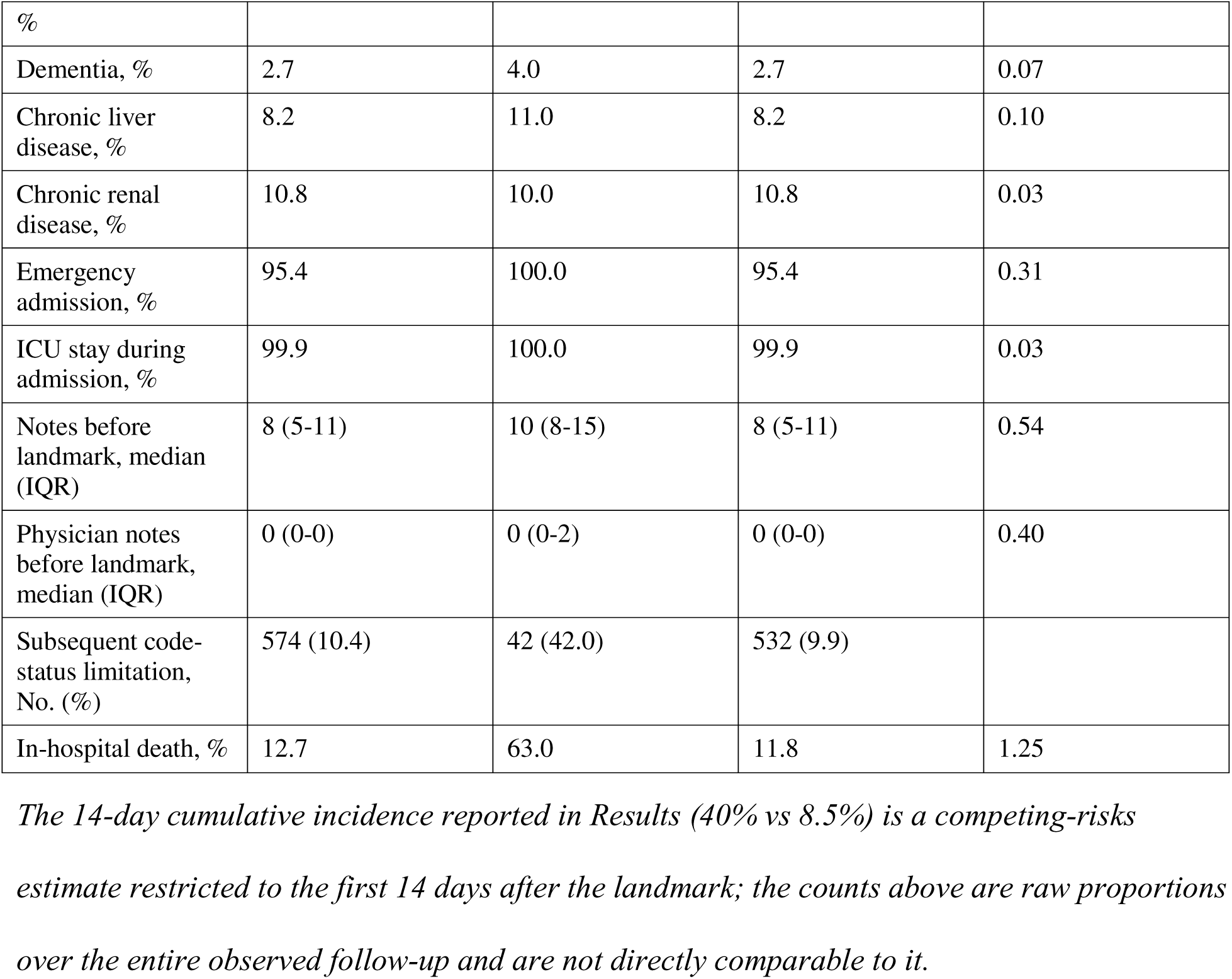
Characteristics of the MIMIC-III 24-hour landmark cohort, by prior documented prognostic language. Covariates are the worst value recorded up to the 24-hour landmark; note counts are documented before the landmark. Continuous medians are computed among observed values, with the percentage missing before the landmark shown for the Glasgow Coma Scale motor score and Richmond Agitation-Sedation Scale. SMD, standardized mean difference; GCS, Glasgow Coma Scale; RASS, Richmond Agitation-Sedation Scale.

### Prognostic language and subsequent code-status limitation (MIMIC-III)

Because MIMIC-IV-Note is not time-stamped, we used MIMIC-III, which retains timestamped notes and code-status orders, to test whether documented prognostic language was associated with a subsequent transition from full code to a code-status limitation, after adjustment for available neurological, comorbidity, and admission characteristics. In a landmark analysis of admissions alive and full code at 24 hours (n = 5,493; 5,171 patients; 100 with prior prognostic language; 574 events; Table 1), prior documented prognostic language was associated with a higher cause-specific hazard of any code-status limitation (adjusted hazard ratio, 4.3; patient-clustered 95% CI, 2.9-6.5; Figure 2). Admissions with prognostic language were sicker (GCS motor 1 vs 5; ventilation 86% vs 50%; mortality 63% vs 12%; Table 1); severity confounding is intrinsic. Adjustment moved the crude estimate (6.7) to 4.3; expanding beyond 5 covariates (3.5) did not reduce it further. The estimate was not materially attenuated after adjustment for acute physiology (laboratory markers and vasopressor exposure; HR, 4.1; 3.6 with missing-data indicators), before-statement covariates (4.6), or documentation-volume adjustment (4.3), and remained elevated in an incidence-density risk-set matched analysis (HR, 5.4; matched-set-stratified, 3.2; eTable 15). The unadjusted 14-day cumulative incidence was 40% versus 8.5%. The association held for the components (comfort measures, HR 3.9 [95% CI, 2.3-6.7]; do-not-resuscitate or do-not-intubate without comfort, HR 4.2 [95% CI, 2.7-6.6]), in a higher-risk neurologic subgroup (HR 4.4 [95% CI, 2.9-6.8]; n = 3,102), and across 48-hour, time-varying, first-admission, and 12-hour-interval analyses (3.3-4.4; Figure 2). Proportional hazards did not hold for the exposure (P = .02): the association was concentrated in the first 3 days (HR, 4.5; 95% CI, 3.0-6.5) and was not statistically distinguishable from the null thereafter (HR, 1.7; 95% CI, 0.9-3.1). Substituting far more common non-prognostic text supported relative specificity: radiographic-severity and diagnostic-hedging language were not associated (HR 1.1 each) and generic critical-illness language only weakly (1.3; Table 2). The association held within both documentation eras and across the hemorrhagic, traumatic, and anoxic-cardiac-arrest phenotypes (eTable 19). The observed-full-code requirement selected a slightly healthier cohort (mortality 12.7% vs 16.8%; eTable 16). Because the same clinical judgment can generate both the statement and the order, this is an association, not evidence of causation (eMethods; eTables 15-20).

**Figure 2.**
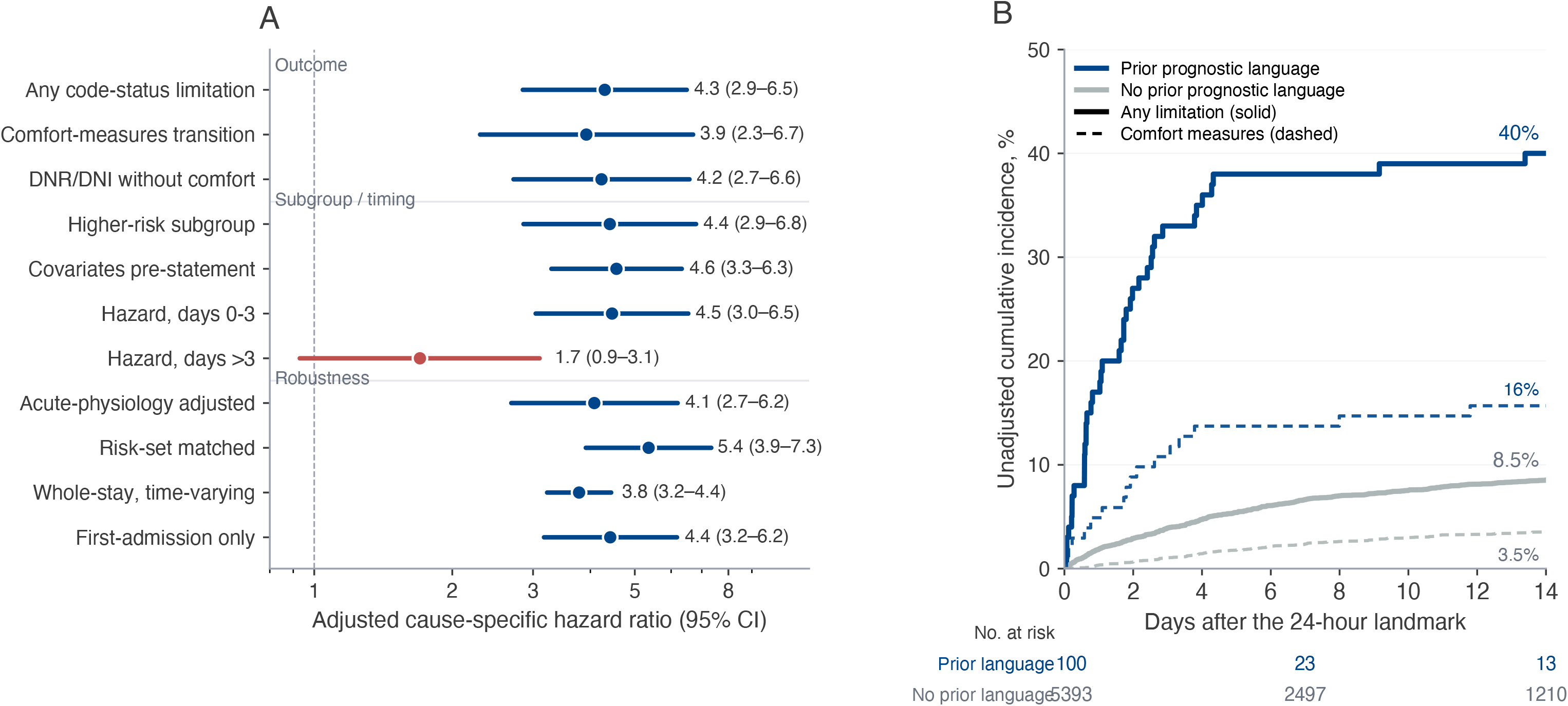
Documented prognostic language and subsequent code-status limitation in the timestamped cohort (MIMIC-III). (A) Adjusted cause-specific hazard ratios (points) with patient-clustered 95% CIs (whiskers; dashed reference at 1) for a subsequent code-status limitation associated with prior documented prognostic language. Rows show the outcome components (any code-status limitation; comfort-measures transition; do-not-resuscitate or do-not-intubate without comfort measures); a higher-risk neurologic subgroup; covariates measured before the statement; the piecewise early (days 0-3) and later (>3 days) hazards, the later estimate (salmon) crossing 1; and further sensitivity analyses (acute-physiology adjustment, incidence-density risk-set matching, whole-stay time-varying, first-admission only). All models adjust for the expanded covariate set (age, phenotype, GCS motor score, ventilation, sedation, van Walraven comorbidity index, metastatic cancer, dementia, chronic liver and renal disease, emergency admission, pre-landmark intensive care unit hours, and database source); death and discharge are competing events. (B) Unadjusted competing-risks cumulative incidence over 14 days after the 24-hour landmark (n = 5,493) for any code-status limitation (solid) and comfort measures (dashed), by prior prognostic language, with numbers at risk below the axis. The association is observational and cannot establish causation.

**Table 2.** Specificity of the code-status association: prognostic language versus non-prognostic text registers (MIMIC-III). Each row is the identical 24-hour landmark cause-specific Cox model (adjusted for the same covariate set; confidence intervals clustered by patient; death and discharge competing), with the exposure replaced by the first documented instance of the register. Registers are mutually contrasted in the extractor: prognostic language is a value judgment about survival or recovery; radiographic severity is objective imaging description; diagnostic-certainty hedging is interpretive uncertainty; generic critical-illness acuity is non-prognostic severity-of-illness language.

| Text register (exposure) | Documented by 24 h, % | Adjusted HR (95% CI) |
| --- | --- | --- |
| Prognostic language | 1.8 | 4.3 (2.8-6.4) |
| Radiographic severity | 83.0 | 1.1 (0.9-1.4) |
| Diagnostic-certainty hedging | 85.4 | 1.1 (0.9-1.4) |
| Generic critical-illness acuity | 16.8 | 1.3 (1.1-1.7) |

*Despite being 40- to 50-fold more common, radiographic-severity and diagnostic-hedging language were not associated with a subsequent code-status limitation (confidence intervals including 1) and generic critical-illness language was only weakly associated (HR 1.3), whereas the rare prognostic-language register showed an approximately fourfold association. This supports relative specificity of the prognostic-language signal rather than an artifact of severity description, interpretive hedging, or documentation density; the registers nonetheless differ in prevalence, timing, and semantics.*

### Temporal ordering of prognosis and responsiveness (MIMIC-III)

In MIMIC-III (11,079 admissions; identical frozen extractors, eTable 14),[26] the timestamped narrative carried the earliest prognostic language a median of 2 days before the discharge summary (first or sole there in 57%). The most recent documented command-following before the first statement was positive in 54.3% but a median of 13 hours old (within 24 hours in only 28%). Among in-hospital deaths, only 29.1% carried any documented prognostic language (eMethods; eTable 14).

### Where prognostic language is written (MIMIC-IV)

Prognostic language was detected in 4.9% (95% CI, 4.6%-5.1%; 1,127 of 23,239) of discharge summaries. Prevalence rose with phenotype lethality, from 1.8% (traumatic brain injury) to 15.4% (cardiac arrest) (Cramér V, 0.20; eFigure 1A); excluding cardiac arrest it was 3.9% to 4.5% (eTable 11).

The extractor detected prognostic language in 27 of 177,855 radiology reports (0.015%; 7 of 41,205 head or brain reports); among admissions with both streams and detected language (n = 1,116), 98.4% localized to the discharge summary alone (eFigure 1B). A targeted audit confirmed that radiologists rarely author prognostic assessments (eTable 4).

### Co-occurrence of prognostic language and documented responsiveness (MIMIC-IV)

Among the 1,127 admissions with prognostic language, 44.4% (95% CI, 41.5%-47.3%) also documented preserved responsiveness at some point and 7.0% near discharge (eTable 6); because the MIMIC-IV narrative is not time-stamped, this co-occurrence is a property of the record, not evidence of responsiveness when a prognosis was formed.

### Completeness of responsiveness documentation (MIMIC-IV and eICU-CRD)

Among the 12,007 admissions with both layers, the discharge summary was silent on command-following in 55.7% (eFigure 2A); without temporal alignment the layers agreed well (Gwet AC1, 0.81; eTable 5), an upper bound because an ever-recorded flag is easily satisfied. A time-matched comparison was limited by coverage: among 698 admissions with an end-of-stay narrative examination (eTable 12), no structured GCS motor score had been charted in the final 24 hours in 508 (72.8%), and among the 190 with a contemporaneous value the layers agreed closely (Gwet AC1, 0.93; eFigure 2B). Within these ICU-derived datasets the narrative examination was frequently the only available record of responsiveness near discharge; this reflects ICU-database coverage, as in eICU-CRD[28] a structured value was absent before hospital discharge in 47.9% of neurologic ICU stays but in only 2.6% before ICU discharge (eTable 13).

### Severe-injury subgroup and construct validity

In the severe-injury subgroup (11,366 admissions), prognostic language was detected in 10.2% (95% CI, 9.6%-10.9%) and every pattern held, so the findings are not an artifact of mild-injury dilution (eTable 2). Structured severity features predicted the presence of prognostic language (AUROC, 0.86) but did not encode the language itself (eTable 7); patient-clustered and first-admission estimates fell within 1.5 percentage points of every descriptive headline (eTable 8).

## Discussion

In this multidatabase study of acute brain injury, prognostic language was written almost entirely in the treating team’s narrative rather than in imaging; command-following was often undocumented and structured charting usually absent near discharge; and, in a timestamped cohort, documented prognostic language was associated with a subsequent code-status limitation after multivariable adjustment. These are measurements of the record, not of clinical truth.

First, prognostic language localizes to the narrative, not the radiology report, so it must be sought there rather than in imaging or structured fields. Second, structured data are an incomplete proxy for responsiveness: the discharge narrative did not mention command-following in more than half of admissions, while the imperfectly reliable structured GCS motor score was frequently unavailable near hospital discharge, so neither layer alone is a complete account.[29–35]

Third, and most consequentially, in the timestamped cohort prior documented prognostic language was associated with a subsequent code-status limitation, an adjusted cause-specific hazard roughly fourfold higher, with an unadjusted 14-day cumulative incidence of 40% versus 8.5%. The association persisted after adjustment for acute physiology, in an incidence-density risk-set match, and across eras and phenotypes, and was largely specific to prognostic language rather than more common non-prognostic text; but residual confounding by evolving illness severity and the clinician’s integrated prognostic judgment, which the measured covariates capture only in part, remains likely, so the analysis cannot distinguish an accurate prognosis appropriately acted upon from a self-fulfilling prophecy.[1–5,36,37] It quantifies at scale a coupling of documented prognosis and subsequent code-status limitation long of concern in neurocritical care. These findings connect computational efforts to surface goals-of-care communication from clinical text[38–43] with neuroprognostication guidelines that caution against premature, self-reinforcing limitation of care,[44–49] and are a reminder that a documented absence of bedside response is an imperfect measure of neurological function.[50–53]

### Limitations

This study has several limitations. First, it is descriptive; the outcome contrasts are confounded by severity by construction. Second, the primary data derive from single-institution databases; the structured end-of-stay finding was echoed across 187 eICU-CRD hospitals but reflects ICU-database coverage ending at ICU transfer, not a validated hospital-wide gap, and the narrative-based analyses could not be validated externally. Third, the MIMIC-IV corpus contains only discharge summaries and radiology reports, so responsiveness and prognosis were measured within two streams, and reported values are detected prevalences. Fourth, the code-status association is observational and derives from a single institution’s older MIMIC-III records (2001-2012), an era predating current neuroprognostication, and MIMIC-III and MIMIC-IV are not independent institutions. Residual confounding by the clinician’s global prognostic judgment, which the available covariates capture only in part despite adjustment for acute physiology (laboratory markers and vasopressor exposure; a full APACHE score and the respiratory organ-failure component were not computable, and sedation scoring was sparse; eTable 18), is the central threat, so a large E-value (eTable 15) does not exclude it. Proportional hazards did not hold for the exposure, so the single hazard ratio summarizes a time-varying effect. A structured code-status order is a proxy for the underlying decision and does not itself establish withdrawal of life-sustaining treatment; whether the documented statement and order reflect a temporal sequence or co-documentation of one conversation cannot be resolved by the association; two physicians agreed on the sequence classification in 61% (Cohen κ, 0.43); among agreements, 52% were a statement preceding a separate later decision and 37% co-documentation of one episode (eTable 21), so even manual review could not reliably distinguish influence from co-documentation. No validated functional outcomes were available to determine whether documented pessimism was accurate or premature. Fifth, the extractors are rule-based, characterized against analyst labels that two physicians confirmed (Cohen κ ≥ 0.96; eTable 4), and remain measurement tools, not instruments validated for individual use. Finally, an unadjusted association with recorded race and ethnicity attenuated after adjustment (eTable 9).

## Conclusions

In this multidatabase study of acute brain injury, prognostic language was written in the treating team’s timestamped narrative rather than in imaging or structured fields, and, in a timestamped cohort, it was associated with a subsequent code-status limitation after adjustment for available neurological, comorbidity, and admission characteristics. This information is distributed across record layers, is largely invisible to structured fields, and is closely coupled with consequential decisions. Testing prospectively whether documented prognostic language shapes subsequent code-status decisions rather than only reflecting severity is a prerequisite for the rigorous study of neuroprognostication and for the responsible secondary use of clinical records.

## Supporting information

Supplementary Information (eMethods, eTables 1-21, eFigures 1-2)

## Data Availability

All data analyzed in the present study are derived from MIMIC-III (version 1.4) and MIMIC-IV (version 3.1), which are publicly available to credentialed users through PhysioNet (https://physionet.org). Access requires completion of the required human-subjects research training and acceptance of the PhysioNet data use agreement. The analysis code is available at https://github.com/Alon-Gorenshtein/Progsontic-unstructured-.

