## Supplementary Information (eMethods, eTables 1-21, eFigures 1-2) for "Prognostic Language and Subsequent Code-Status Limitation After Acute Brain Injury: A Multidatabase Observational Study"

This supplement contains eMethods (cohort and extractor detail, timing definitions, statistics, a model card, and the MIMIC-III companion and code-status limitation analyses), eTables 1 to 21, and eFigures 1 to 2.

### eMethods

#### 1. Cohort and phenotype definitions

Admissions were drawn from MIMIC-IV version 3.1 and linked to MIMIC-IV-Note version 2.2 by subject_id and hadm_id. The linked corpus contains discharge summaries and radiology reports only; it holds no progress, consultation, or goals-of-care notes, and all text-based measures are scoped to these two streams. Acute brain injury was defined by International Classification of Diseases, Ninth and Tenth Revision (ICD-9 and ICD-10) codes (eTable 1). When more than 1 phenotype was coded for an admission, a single primary phenotype was assigned by a fixed acuity priority (cardiac arrest, anoxic-ischemic injury, subarachnoid hemorrhage, intracerebral hemorrhage, traumatic brain injury, subdural hemorrhage, acute ischemic stroke, status epilepticus, central nervous system infection), and all co-occurring phenotypes were retained as flags for sensitivity analysis (eTable 10). Aim-specific denominators were: admissions with a discharge summary or a head-imaging report (Aim 1), admissions with both a discharge summary and structured Glasgow Coma Scale (GCS) motor charting (Aim 2), and admissions with a discharge summary (Aim 3). A secondary, clinically defined severe-injury subgroup comprised admissions with an intensive care unit (ICU) stay together with either mechanical ventilation or a hemorrhagic, anoxic-ischemic, or cardiac-arrest phenotype (11,366 admissions; 28.7% in-hospital mortality; 8,700 with a discharge summary). The severe-injury subgroup is reported as a sensitivity analysis; the full cohort is the primary descriptive sample.

#### 2. Prognostic-language extractor

Prognostic language was defined as a value-laden statement about a patient's expected survival or neurological recovery. The rule-based extractor captured seven prognostic-language groups: poor prognosis ("poor/grave/grim/guarded/dismal prognosis," including forms separated by intervening words such as "prognosis was extremely poor"), devastation ("devastating brain injury," "catastrophic brain injury"), nonsurvivability ("nonsurvivable," "incompatible with life"), absent meaningful recovery ("unlikely to have meaningful recovery," "no meaningful recovery"), unsalvageability ("no salvageable brain"), futility ("futile," "medically futile"), and terminal framing ("not expected to survive," "expected to die"). Two contrasting registers were measured to establish that prognostic language is a distinct construct: radiographic severity ("midline shift," "herniation," "mass effect," "large hemorrhage") and diagnostic-certainty hedging ("consistent with," "cannot exclude," "concerning for").

Five guards encoded the adjudication rubric to exclude non-prognostic matches: (1) procedural futility (eg, "further attempts were futile" during a catheter procedure); (2) negated prognosis ("without evidence to indicate poor prognosis"); (3) organ-specific recovery ("recovery of vision"); (4) a family member as the grammatical subject ("his sister had a devastating stroke"); and (5) conditional or negated valence ("if prognosis was poor," "prognosis is not poor"). The extractor was frozen before evaluation. Radiology-report matches were localized by report section (indication or clinical history vs findings or impression) to separate language authored by the referring clinician from language authored by the radiologist.

#### 3. Responsiveness documentation and timing definitions

Narrative command-following was extracted from discharge summaries with negation-aware logic: mentions of following or obeying commands were affirmative unless preceded within a short window by a negation cue ("not," "unable to," "does not," "no," "never," "cannot"). Each summary was classified as documenting command-following, its explicit absence, both, or neither. Narrative purposeful responsiveness captured visual pursuit, tracking, and localization to stimulus. Documented command-following is a sign of behavioral responsiveness; it is not read here as a measure of level of awareness, which can be retained without any observable behavioral response.

Structured command-following was a GCS motor score of 6 ("obeys commands," item 223901). Three timing definitions were used:

- **Any-time (unaligned):** command-following recorded at any point during the admission (structured) or stated anywhere in the discharge summary (narrative). Used for the stay-level agreement upper bound and the any-time co-occurrence.
- **Final 24 hours (structured):** a GCS motor score of 6 recorded within 24 hours before the terminal event (death or discharge).
- **End-of-stay narrative examination:** command status parsed from the physical-examination section of the discharge summary.

The time-matched comparison paired the end-of-stay narrative examination with the final-24-hour structured measure, restricted to admissions in which the narrative examination gave an unambiguous positive or negative status (n = 698 full; n = 475 severe). "Late" responsiveness for the co-occurrence analysis required either the final-24-hour structured measure or the end-of-stay narrative examination to be positive.

#### 4. Structured extraction

Structured signals were streamed from chartevents (approximately 330 million rows) with a memory-efficient filter restricted to the cohort and to five item identifiers: GCS eye (220739), verbal (223900), and motor (223901); Richmond Agitation-Sedation Scale (228096); and code status (223758). Per admission, we recorded the best (maximum) motor score, whether command-following was ever recorded, the final-24-hour motor score, the minimum sedation score, and code-status transitions.

#### 5. Statistical analysis and model card

Prevalences are reported with Wilson 95% CIs and, because patients could contribute more than 1 admission, with patient-clustered bootstrap CIs (2,000 resamples of patients; seed, 20260719) and a first-admission-only sensitivity analysis (eTable 8). Between-phenotype differences used the χ^2^ test with Cramér V. Agreement used the Gwet AC1 statistic and Cohen κ, with the McNemar test for directional differences. The Spearman correlation summarized the phenotype-level relationship between detected prognostic-language prevalence and mortality. Multiplicity across the primary family of tests was controlled with the Benjamini-Hochberg false-discovery-rate procedure (2-sided P < .05).

**Model card (construct-validity probe).** Purpose: to test how well structured severity features identify admissions containing prognostic language (a construct-validity probe, not a reducibility test). Models: L2-regularized logistic regression (standardized features) with isotonic calibration, and histogram gradient boosting. Features: primary phenotype (one-hot), age, sex, best GCS motor, ever-obeys flag, minimum sedation score, ICU admission, and length of stay. Target: presence of narrative prognostic language. Training and evaluation: 5-fold stratified cross-validation with out-of-fold predictions; AUROC with 2,000-sample bootstrap CIs and Brier score; models compared by paired bootstrap. Result: AUROC 0.858 (logistic) and 0.859 (gradient boosting); Brier 0.038 for both (eTable 7). Intended use: measurement only. This is not a deployable clinical tool, was not externally validated, and must not be used to identify patients for care decisions.

**Multicenter database-coverage analysis (eICU-CRD).** Because the end-of-stay structured-documentation pattern could reflect either genuine undercharting or the coverage boundary of an intensive care database, we examined it across multiple centers using the eICU Collaborative Research Database version 2.0, a deidentified multicenter database of intensive care admissions to approximately 200 US hospitals during 2014 to 2015.[28] This is a coverage check, not an external validation of the narrative-based analyses. We defined a neurologic cohort from the APACHE admission diagnosis (cerebrovascular accident, intracranial hemorrhage, subarachnoid hemorrhage, head trauma, cardiac arrest, anoxic injury, seizure or status epilepticus, coma or altered level of consciousness, encephalopathy, and central nervous system infection). Structured neurological documentation was a charted Glasgow Coma Scale or motor-response value in the nurse-charting table. For each stay with such charting, we computed whether a value was recorded within 24 hours before hospital discharge or death (comparable to the MIMIC-IV final-24-hour measure) and, as a mechanism check, within 24 hours before intensive care unit discharge. Results are reported overall, by hospital-discharge status, and as the distribution across hospitals with at least 20 qualifying stays.

**Timestamped-note companion analysis (MIMIC-III).** Because MIMIC-IV-Note contains only discharge summaries and radiology reports, the temporal ordering of prognostic language could not be examined in the primary corpus. We therefore repeated the extraction in MIMIC-III version 1.4, a deidentified critical-care database from the same institution covering 2001 to 2012 that retains timestamped physician, nursing, consultation, and progress notes.[26] We assembled an acute brain injury cohort using the same ICD-9 phenotype families (11,079 admissions; 9,794 patients; 98.6% with an intensive care unit stay; 18.3% in-hospital mortality) and applied the identical, frozen prognostic-language and command-following extractors, adding a goals-of-care register split into code status (do-not-resuscitate or do-not-intubate) and active limitation (comfort care, withdrawal of life-sustaining treatment, or palliation). Each note was placed on the admission timeline by its charted time; 344,554 of 387,975 cohort notes (88.8%) carried a minute-resolution timestamp, and primary temporal analyses were restricted to these. We measured, per admission, the timing and note category of the first prognostic-language statement (Aim 1); the most recent command-following documentation at or before that statement (Aim 2); and the ordering of the statement relative to the first active-limitation note and to in-hospital death (Aim 3). Proportions are summarized with Wilson and patient-clustered bootstrap 95% CIs. Because MIMIC-III derives from the same institution and an overlapping era, this analysis examines temporal ordering rather than providing independent replication, and the notes remain documentation rather than the underlying clinical events.

**Code-status limitation association (MIMIC-III).** Because MIMIC-III also records timestamped code-status orders, we tested whether documented prognostic language was associated with a subsequent structured transition from full code to a code-status limitation, with adjustment for available measures of severity and comorbidity. The exposure was the first documented prognostic-language statement (physician-adjudicated register; eTable 14); the outcome was the first structured code-status transition recorded in chartevents, using the reliable structured field rather than the lower-precision note-based active-limitation register (eTable 14). Because these decisions differ clinically, we analyzed the outcome as a composite (any of do-not-resuscitate, do-not-intubate, or comfort measures) and by component: comfort-measures transitions, and do-not-resuscitate or do-not-intubate orders without comfort measures. The composite risk set excludes admissions with any limitation by the landmark, whereas the comfort risk set excludes only those with a prior comfort transition, so an admission that reached do-not-resuscitate or do-not-intubate (without comfort) by the landmark remains at risk for a subsequent comfort transition and enlarges the comfort risk set (5,512 vs 5,493 at 24 hours; the 19 additional admissions had a do-not-resuscitate or do-not-intubate order without comfort by 24 hours). The comfort model therefore estimates the cause-specific hazard of any subsequent full-code-to-comfort transition, treating do-not-resuscitate or do-not-intubate as an intermediate rather than a censoring state, with death and discharge competing. The primary analysis was a cause-specific Cox model at 24- and 48-hour landmarks, restricted to admissions alive, still admitted, and whose most recent observed code-status state before the landmark was full code (default full code was not assumed; admissions with no observed code-status entry before the landmark were excluded). Because eligibility requires an observed full-code entry at or before the landmark, and some admissions record their first code-status entry between 24 and 48 hours, the 48-hour risk set is not nested in the 24-hour set. Death and discharge were competing events. To address severity confounding, the adjustment set was expanded from 5 covariates to age, primary phenotype, worst Glasgow Coma Scale motor score, mechanical ventilation, and deepest Richmond Agitation-Sedation Scale value (all measured up to the landmark), the van Walraven comorbidity index (Quan 2005 International Classification of Diseases, Ninth Revision map), metastatic cancer, dementia, chronic liver disease, chronic renal disease, emergency admission, intensive care unit hours elapsed to the landmark, and database source (CareVue or MetaVision). Intensive care unit hours are counted only up to the landmark; total intensive care unit length of stay and the number of intensive care unit stays were deliberately excluded because they are determined partly after the exposure and the outcome (post-outcome colliders). A further model adjusted for acute physiology, using the MIMIC-III laboratory and infusion tables: the worst value up to the landmark for serum creatinine, total bilirubin, platelet count, lactate, blood urea nitrogen, and arterial oxygen tension (labevents), and any vasopressor exposure before the landmark (norepinephrine, epinephrine, dopamine, dobutamine, phenylephrine, or vasopressin from inputevents_cv and inputevents_mv, using the standard CareVue and MetaVision item identifiers). Missing laboratory values were set to the cohort median. A full APACHE score and the respiratory Sequential Organ Failure Assessment component were not computed because fraction-of-inspired-oxygen charting was too sparse to form reliable ratios; this residual gap is a limitation.

We also performed an incidence-density risk-set matched analysis: for each first prognostic-language statement, up to 5 controls were sampled at random from admissions alive, admitted, full-code, and limitation-free and not yet exposed at the statement hour (controls were allowed to become exposed later, and were censored at their own first statement, so that no post-index information defined the control set); the statement time was the index for the case and its controls; covariates (including acute physiology) were measured only before that index; and the cause-specific hazard was estimated from the index time, clustered by patient, with a matched-set-stratified model as a sensitivity. As a specificity test, the exposure was replaced by non-prognostic text registers detected by the same frozen extractor logic (radiographic severity, eg "midline shift," "large hemorrhage"; diagnostic-certainty hedging, eg "consistent with," "cannot exclude"; and a generic critical-illness/acuity register, eg "critically ill," "hemodynamically unstable," "deteriorating"), each entered as the first-occurrence exposure in the identical landmark model (Table 2; eTable 20). We report the primary association stratified by documentation era (CareVue, MetaVision) and by phenotype (eTable 19). Finally, to probe whether the statement-to-order pairing is a temporal sequence or co-documentation of a single conversation, two physicians independently and blindly classified the 143 admissions with a code-status limitation within 48 hours after the first statement into one of seven categories (statement preceded a separate later decision; same goals-of-care episode; prior directive or family preference; retrospective documentation; copied-forward language; clinical deterioration between statement and order; or timing/relationship unclear); inter-rater agreement was summarized with raw agreement and Cohen κ (eTable 21). Models were unpenalized and reported with confidence intervals clustered by patient; proportional hazards were tested with scaled Schoenfeld residuals, and because the assumption did not hold for the exposure we report a piecewise hazard ratio split at 72 hours (a small ridge penalty of 0.1 was used only for the piecewise fits, which have sparse events). Because landmark covariates measured "up to the landmark" can post-date an early statement, a sensitivity analysis restricted covariates for exposed admissions to values recorded strictly before the first statement (and for unexposed admissions before the landmark). Because that pre-statement window is shorter for admissions with an early statement (an asymmetry between the exposed and unexposed groups), we also performed an equal-covariate-window analysis: each unexposed admission was assigned a pseudo-index time drawn at random (seed 1) from the exposed statement-time distribution, both groups were required to be alive, admitted, full-code, and limitation-free at their own index, and covariates for both groups were measured only before their own index; the cause-specific hazard was then estimated from the index time. A secondary whole-stay time-varying Cox model let the exposure become active at the first statement (avoiding immortal-time bias) with baseline (first-24-hour and fixed) covariates rather than worst-over-stay values. Additional analyses were a first-admission-only sensitivity analysis, a required 12-hour statement-to-order interval, the timing distribution between statement and order, the association within a higher-risk neurologic subgroup (a hemorrhagic [intracerebral, subarachnoid, or subdural], anoxic-ischemic, or cardiac-arrest phenotype, or any mechanical ventilation), a model further adjusted for pre-landmark documentation volume (the natural logarithm of the total and physician note counts documented before the landmark), a characterization of admissions included versus excluded by the observed-full-code requirement (eTable 16), the covariate completeness of the Glasgow Coma Scale motor score, Richmond Agitation-Sedation Scale value, and ventilation by exposure group (eTable 18), the unadjusted competing-risks cumulative incidence at 14 days for the composite and comfort-measures outcomes, and an E-value.[27] We also report the association with the competing outcome of discharge alive; because a genuine poor prognosis is itself expected to reduce discharge alive, this is a descriptive competing-outcome contrast rather than a strong falsification test. Because the same clinical judgment can generate both the written prognosis and the order, this association is not evidence of causation.

Software: Python 3.9 with pandas, NumPy, SciPy, scikit-learn, statsmodels, and lifelines.

### eTables

#### eTable 1. Acute brain injury phenotype definitions (ICD-9 and ICD-10 code prefixes)

| Phenotype | ICD-9 | ICD-10 |
| --- | --- | --- |
| Cardiac arrest | 427.5 | I46 |
| Anoxic-ischemic injury | 348.1, 348.5 | G93.1, G93.82 |
| Subarachnoid hemorrhage | 430 | I60 |
| Intracerebral hemorrhage | 431 | I61 |
| Subdural hemorrhage | 432 | I62 |
| Acute ischemic stroke | 433, 434 | I63 |
| Traumatic brain injury | 800-804, 850-854 | S06 |
| Status epilepticus | 345.3 | G41 |
| Central nervous system infection | 320-322 | G00, G03 |

#### eTable 2. Detected prognostic-language prevalence in discharge summaries, by phenotype (full cohort and severe subgroup)

| Phenotype | Full: admissions | Full: with language, No. | Full: detected prevalence, % (95% CI) | Severe: admissions | Severe: with language, No. | Severe: detected prevalence, % (95% CI) |
| --- | --- | --- | --- | --- | --- | --- |
| Cardiac arrest | 1,846 | 284 | 15.4 (13.8-17.1) | 1,637 | 274 | 16.7 (15.0-18.6) |
| Anoxic-ischemic injury | 2,922 | 305 | 10.4 (9.4-11.6) | 1,877 | 268 | 14.3 (12.8-15.9) |
| Intracerebral hemorrhage | 2,242 | 166 | 7.4 (6.4-8.6) | 1,571 | 129 | 8.2 (7.0-9.7) |
| Subdural hemorrhage | 916 | 48 | 5.2 (4.0-6.9) | 517 | 36 | 7.0 (5.1-9.5) |
| Subarachnoid hemorrhage | 935 | 48 | 5.1 (3.9-6.7) | 756 | 43 | 5.7 (4.3-7.6) |
| Status epilepticus | 252 | 11 | 4.4 (2.5-7.7) | 142 | 10 | 7.0 (3.9-12.5) |
| Central nervous system infection | 413 | 9 | 2.2 (1.2-4.1) | 67 | 5 | 7.5 (3.2-16.3) |
| Acute ischemic stroke | 7,835 | 148 | 1.9 (1.6-2.2) | 1,089 | 65 | 6.0 (4.7-7.5) |
| Traumatic brain injury | 5,878 | 108 | 1.8 (1.5-2.2) | 1,044 | 59 | 5.7 (4.4-7.2) |
| **Overall** | **23,239** | **1,127** | **4.9 (4.6-5.1)** | **8,700** | **889** | **10.2 (9.6-10.9)** |

Full cohort: χ^2^ across phenotypes P < .001; Cramér V, 0.20; Spearman ρ between phenotype detected prevalence and mortality, 0.92. Severe subgroup: χ^2^ P < .001; Cramér V, 0.15.

#### eTable 3. Prognostic-language groups (full cohort)

| Prognostic-language group | Language-positive summaries containing group, No. (%) |
| --- | --- |
| Poor prognosis | 954 (84.6) |
| Futility | 80 (7.1) |
| Absent meaningful recovery | 60 (5.3) |
| Devastation | 59 (5.2) |
| Nonsurvivability | 31 (2.8) |
| Terminal framing | 18 (1.6) |
| Unsalvageability | 3 (0.3) |

Groups are not mutually exclusive; a summary may contain more than one. Denominator, 1,127 language-positive discharge summaries.

#### eTable 4. Operating characteristics of the two extractors against analyst and two-physician consensus reference labels

Reference labels were adjudicated by a single analyst against a written rubric on stratified random samples of discharge summaries (a detected pool, a random not-detected pool, and a not-detected pool enriched for likely misses). The random not-detected pool gives an unbiased 2×2 against the analyst reference; the design-weighted columns correct for the sampling fractions to estimate population-level precision, recall, specificity, and F1. Two physicians independently adjudicated the same excerpts, and their consensus is reported here as a second, independent reference standard (see the two-physician adjudication table below for the underlying agreement statistics). Each design-weighted column reports precision, recall, specificity, and F1 against a single named reference, so no recall estimate is paired with a specificity computed against a different reference or sample.

**Prognostic-language extractor**

| Metric | Random-negative pool (raw 2×2, vs analyst) | Design-weighted vs analyst reference | Design-weighted vs two-physician consensus |
| --- | --- | --- | --- |
| 2×2 counts (TP / FP / FN / TN) | 75 / 5 / 1 / 119 | NA | NA |
| Precision (PPV) | 0.94 (95% CI, 0.86-0.97) | 0.94 (95% CI, 0.88-0.99) | 0.91 |
| Recall (sensitivity) | 0.99 (95% CI, 0.93-1.00) | 0.85 (95% CI, 0.65-1.00) | 0.85 |
| Specificity | 0.96 (95% CI, 0.91-0.98) | 0.997 (95% CI, 0.994-0.999) | 0.996 |
| F1 | 0.96 (95% CI, 0.93-0.99) | 0.89 (95% CI, 0.75-0.99) | 0.88 |

**Command-following extractor**

| Metric | Random-negative pool (raw 2×2, vs analyst) | Design-weighted vs analyst reference | Design-weighted vs two-physician consensus |
| --- | --- | --- | --- |
| 2×2 counts (TP / FP / FN / TN) | 48 / 2 / 6 / 114 | NA | NA |
| Precision (PPV) | 0.96 (95% CI, 0.87-0.99) | 0.96 (95% CI, 0.90-1.00) | 0.96 |
| Recall (sensitivity) | 0.89 (95% CI, 0.78-0.95) | 0.91 (95% CI, 0.85-0.97) | 0.91 |
| Specificity | 0.98 (95% CI, 0.94-1.00) | 0.98 (95% CI, 0.95-1.00) | 0.98 |
| F1 | 0.92 (95% CI, 0.86-0.97) | 0.93 (95% CI, 0.89-0.98) | 0.94 |

Operating characteristics are design-weighted to the source population and computed against the two-physician consensus reference; command-following was identical against the analyst reference, and prognostic-language precision was 0.94 against the analyst reference.

**Second-annotator agreement (automated model, not a clinician).** A locally run Gemma 2 9B model (temperature 0) relabeled the same excerpts from a fixed yes/no rubric.

| Extractor | n | Percent agreement | Cohen κ (95% CI) | Gwet AC1 (95% CI) |
| --- | --- | --- | --- | --- |
| Prognostic language | 280 | 93.6 | 0.85 (0.78-0.91) | 0.89 (0.83-0.94) |
| Command-following | 200 | 98.5 | 0.96 (0.91-1.00) | 0.98 (0.94-1.00) |

Residual prognostic-language false positives were confined to irrealis-mood statements ("would not want futile care," "could have been a devastating injury"). Because prognostic language is rare, reported values are detected prevalences and may be affected by imperfect sensitivity and precision. Radiology-report prognostic language, by section: indication or clinical history, 63%; impression or findings, 26%; other (technique or comparison), 11% (n = 27).

In a targeted recall audit, 88 radiology reports not flagged by the extractor contained a high-risk prognostic term (prognosis, devastating, nonsurvivable, futile, expected to die, incompatible with life, poor outcome, catastrophic); 76 carried the term in the referring clinician's indication or history line and 10 in a radiologist-authored section, and manual review of those 10 found none to be a patient-level prognostic assessment (they were radiographic correlations, negated findings, or lesion-specific statements). Two reports fell in other sections. This supports that radiologists rarely author prognostic assessments and that the low radiology detection rate is not primarily a recall artifact.

**Independent two-physician adjudication.** Two physicians independently relabeled the full set of adjudication excerpts (280 prognostic-language, 200 command-following), each blinded to the extractor output, to the analyst reference labels, and to the other physician's judgments.

| Comparison | Prognostic language: κ (AC1) | Command-following: κ (AC1) |
| --- | --- | --- |
| Physician 1 vs analyst reference | 0.96 (0.98) | 1.00 (1.00) |
| Physician 2 vs analyst reference | 0.96 (0.98) | 1.00 (1.00) |
| Physician 1 vs physician 2 | 0.98 (0.99) | 1.00 (1.00) |
| Human consensus vs automated extractor | 0.92 (0.95) | 0.88 (0.93) |

The two physicians agreed with each other and with the analyst reference labels on 278 of 280 prognostic-language excerpts (99.3%) and on all 200 command-following excerpts (100%). This confirms the reference labels used for the operating-characteristic estimates and establishes that the extractor definitions correspond to clinician judgment.

#### eTable 5. Responsiveness documentation and structured-vs-narrative agreement (admissions with both layers)

| Measure | Full (n = 12,007) | Severe (n = 8,653) |
| --- | --- | --- |
| Narrative documents command-following (positive only), % | 30.8 | 28.3 |
| Narrative documents explicit absence (negative only), % | 7.3 | 8.5 |
| Narrative documents both (mixed), % | 6.2 | 6.7 |
| Narrative silent, % | 55.7 | 56.5 |
| Structured GCS motor ever "obeys commands," % | 85.6 | 81.5 |
| Unaligned stay-level agreement (n = 4,573 full; 3,183 severe): Gwet AC1 | 0.81 | 0.76 |
| Unaligned stay-level agreement: Cohen κ | 0.44 | 0.44 |
| Final-24h GCS motor charted (coverage), n / N (%) | 190 / 698 (27.2) | 119 / 475 (25.1) |
| - No final-24h GCS motor score charted, n (%) | 508 (72.8) | 356 (74.9) |
| Agreement among admissions with a final-24h value (n = 190 full; 119 severe): Gwet AC1 | 0.93 | 0.92 |
| Agreement among admissions with a final-24h value: Cohen κ | 0.47 | 0.52 |
| Agreement among admissions with a final-24h value: raw, % | 93.7 | 93.3 |

The four narrative categories are mutually exclusive and exhaustive. Unaligned comparison: any clear narrative statement vs structured command-following ever recorded during the stay (n = 4,573 full; 3,183 severe). Time-matched comparison: end-of-stay narrative examination vs structured GCS in the final 24 hours; a structured GCS motor score had been charted in the final 24 hours in only 190 of 698 admissions (27.2%). Among those with a contemporaneous value the layers agreed closely (Gwet AC1, 0.93; 93.7% raw). The absence of a final-24-hour GCS motor score is missing data, not negative clinical evidence, so it is excluded from the agreement statistic and reported separately as coverage.

#### eTable 6. Co-occurrence of prognostic language and preserved responsiveness (admissions with a discharge summary)

| Group | Full: No. | Full: co-occurrence, % (95% CI) | Severe: No. | Severe: co-occurrence, % (95% CI) |
| --- | --- | --- | --- | --- |
| With prognostic language | 1,127 | NA | 889 | NA |
| Any-time preserved responsiveness | 500 | 44.4 (41.5-47.3) | 409 | 46.0 (42.8-49.3) |
| Late preserved responsiveness | 79 | 7.0 (5.7-8.7) | 70 | 7.9 (6.3-9.8) |
| Structured command-following (ever) | 360 | 31.9 (29.3-34.7) | 310 | 34.9 (31.8-38.1) |
| Narrative command-following | 186 | 16.5 (14.5-18.8) | 132 | 14.9 (12.7-17.3) |

Any-time preserved responsiveness by high-lethality phenotype (full cohort): cardiac arrest 41.5%, anoxic-ischemic injury 40.3%, intracerebral hemorrhage 37.3%, subarachnoid hemorrhage 43.8%. In-hospital mortality among admissions with prognostic language was 72.4% (full) and 80.9% (severe); these outcome figures are descriptive and confounded by severity by construction. The co-occurrence is documentary (over the admission), not contemporaneous: responsiveness earlier in a stay is compatible with a later, well-founded poor prognosis, and the narrative prognostic language is not time-stamped.

#### eTable 7. Construct-validity probe and calibration

| Model | AUROC (95% CI) | Brier score | Calibration |
| --- | --- | --- | --- |
| Logistic regression | 0.858 (0.846-0.870) | 0.038 | Isotonic (nested within each out-of-fold split) |
| Gradient boosting | 0.859 (0.847-0.871) | 0.038 | None (raw probabilities) |

Paired bootstrap difference (gradient boosting minus logistic), 0.002 (95% CI, -0.002 to 0.006; P = .37). Strongest logistic predictors (standardized coefficients): ICU admission (+0.98), absence of structured command-following (-0.93 on the ever-obeys flag), deeper sedation (-0.31), older age (+0.24), anoxic-ischemic (+0.24) and cardiac-arrest (+0.17) phenotype. Calibration was corrected from an earlier uncalibrated logistic fit (Brier, 0.14) to 0.038 by isotonic calibration.

**Reliability of the calibrated logistic model (deciles of predicted probability).**

| Decile | Mean predicted | Observed fraction |
| --- | --- | --- |
| 1 | 0.001 | 0.003 |
| 2 | 0.004 | 0.006 |
| 3 | 0.006 | 0.007 |
| 4 | 0.009 | 0.008 |
| 5 | 0.012 | 0.011 |
| 6 | 0.022 | 0.024 |
| 7 | 0.033 | 0.038 |
| 8 | 0.040 | 0.035 |
| 9 | 0.071 | 0.073 |
| 10 | 0.290 | 0.283 |

Predicted and observed frequencies agree across the range, indicating good calibration. The model shows that structured severity features identify admissions in which prognostic language is more likely to appear; they do not encode the language or its content.

#### eTable 8. Patient-clustered and first-admission sensitivity analyses

| Quantity | Naive Wilson, % (95% CI) | Patient-clustered bootstrap, % (95% CI) | First-admission only, % | Δ vs main, pts |
| --- | --- | --- | --- | --- |
| Detected prognostic-language prevalence, full | 4.85 (4.58-5.13) | 4.85 (4.57-5.12) | 5.07 | 0.22 |
| Detected prognostic-language prevalence, severe | 10.22 (9.60-10.87) | 10.22 (9.60-10.85) | 10.37 | 0.15 |
| Narrative silence, full | 55.73 (54.84-56.61) | 55.73 (54.83-56.67) | 55.61 | 0.12 |
| Narrative silence, severe | 56.49 (55.44-57.53) | 56.49 (55.45-57.50) | 56.59 | 0.10 |
| Co-occurrence any-time, full | 44.37 (41.49-47.28) | 44.37 (41.54-47.12) | 43.61 | 0.76 |
| Co-occurrence any-time, severe | 46.01 (42.75-49.29) | 46.01 (42.65-49.44) | 44.58 | 1.43 |
| Co-occurrence late, full | 7.01 (5.66-8.65) | 7.01 (5.55-8.53) | 6.65 | 0.36 |
| Co-occurrence late, severe | 7.87 (6.28-9.83) | 7.87 (6.08-9.82) | 7.60 | 0.27 |

Patient-clustered CIs (2,000 patient resamples; 19,762 patients contributed the 23,239 discharge summaries in the full cohort) are close to the naive intervals. First-admission-only estimates, which remove repeated admissions entirely, fall within 1.5 percentage points of the main estimates for every quantity.

#### eTable 9. Race and ethnicity distribution and adjusted odds of prognostic language (exploratory)

| Race/ethnicity | N (%) of cohort | Detected prevalence, % | Unadjusted OR (95% CI) | Adjusted OR (95% CI) | P (adjusted) |
| --- | --- | --- | --- | --- | --- |
| White (reference) | 16,308 (70.2) | 3.8 | 1.00 (ref) | 1.00 (ref) | NA |
| Black | 2,225 (9.6) | 4.5 | 1.20 (0.97-1.48) | 1.04 (0.83-1.30) | .74 |
| Hispanic/Latino | 826 (3.6) | 4.1 | 1.08 (0.76-1.54) | 1.04 (0.72-1.50) | .84 |
| Asian | 643 (2.8) | 7.1 | 1.94 (1.42-2.65) | 1.47 (1.06-2.05) | .02 |
| Unknown/unspecified/other | 3,237 (13.9) | 10.0 | 2.79 (2.43-3.21) | 1.65 (1.42-1.92) | <.001 |

Cohort N = 23,239 (discharge summary and non-missing age). Overall 70.2% of the cohort is recorded as White versus 55.3% of language-positive records; the unadjusted gap attenuates after adjustment for age, sex, primary phenotype, ICU admission, and any low motor score. "Unknown/unspecified/other" combines patients whose recorded race/ethnicity was not White, Black, Hispanic/Latino, or Asian (including unknown, unable to obtain, declined to answer, and other or multiple categories). This is an exploratory cross-sectional association, not a causal claim; residual confounding by unmeasured severity or documentation practice cannot be excluded.

#### eTable 10. Phenotype-hierarchy assignment and co-occurring flags (sensitivity)

A single primary phenotype was assigned by the fixed acuity priority below; all co-occurring qualifying phenotypes were retained as flags in the analytic dataset.

| Priority | Phenotype |
| --- | --- |
| 1 | Cardiac arrest |
| 2 | Anoxic-ischemic injury |
| 3 | Subarachnoid hemorrhage |
| 4 | Intracerebral hemorrhage |
| 5 | Traumatic brain injury |
| 6 | Subdural hemorrhage |
| 7 | Acute ischemic stroke |
| 8 | Status epilepticus |
| 9 | Central nervous system infection |

Because overlapping diagnoses were assigned using a fixed hierarchy, phenotype-specific estimates should be interpreted as estimates under this classification scheme, and co-occurring-phenotype flags were retained for alternative specifications. The phenotype-level Spearman correlation (ρ, 0.92) rests on 9 phenotype observations and is reported descriptively.

#### eTable 11. Cohort-definition sensitivity: detected prognostic-language prevalence by inclusion criterion

The cohort comprises acute brain injury and related acute neurologic emergencies. Cardiac arrest, status epilepticus, and central nervous system infection were included as emergencies that frequently cause or place the brain at risk of injury. To test whether the headline detected prevalence depends on the broadest inclusions, we recomputed it after removing cardiac-arrest admissions.

| Cohort definition | Discharge summaries, No. | Detected prognostic language, No. (%) | 95% CI |
| --- | --- | --- | --- |
| Full cohort (primary) | 23,239 | 1,127 (4.85) | 4.58-5.13 |
| Excluding cardiac-arrest admissions without a concomitant anoxic-ischemic diagnosis | 21,725 | 968 (4.46) | 4.19-4.74 |
| Excluding all cardiac-arrest-primary admissions | 21,393 | 843 (3.94) | 3.69-4.21 |

Detected prevalence and the phenotype gradient are stable across cohort definitions; the low imaging rate and the documentation-coverage findings do not depend on including cardiac-arrest admissions. Head or brain imaging reports carried prognostic language at a rate (7 of 41,205; 0.017%) similar to that in all radiology reports (27 of 177,855; 0.015%), so the low imaging rate is not an artifact of including non-neurologic studies in the denominator.

#### eTable 12. Time-matched analysis subset versus other both-layer admissions (selection check)

The time-matched comparison (Aim 2) is restricted to admissions with an interpretable end-of-stay narrative examination. This table compares those admissions with the remaining admissions that had both a discharge summary and structured motor charting.

| Characteristic | End-of-stay-examined (n = 698) | Other both-layer (n = 11,309) |
| --- | --- | --- |
| Median age, y | 68 | 68 |
| In-hospital mortality, % | 0.6 | 21.8 |
| ICU admission, % | 100 | 100 |
| Mechanically ventilated, % | 38.4 | 49.9 |
| Severe-injury subgroup, % | 68.1 | 72.3 |
| Median hospital length of stay, d | 7.9 | 7.2 |

The end-of-stay-examined subset is survivor-enriched (in-hospital mortality 0.6% vs 21.8%), consistent with requiring survival to a documented discharge examination. The time-matched result therefore describes a selected subset and should not be read as representative of the full cohort.

#### eTable 13. Multicenter database-coverage analysis in eICU-CRD: structured neurological documentation near end of stay (187 hospitals)

Neurologic intensive care unit stays with structured Glasgow Coma Scale or motor-response charting (n = 26,354) across 187 hospitals.

| Measure | Value |
| --- | --- |
| No structured GCS or motor value in the final 24 h before **hospital** discharge or death, % | 47.9 |
| Absent among patients discharged alive, % | 53.0 |
| Absent among patients who died, % | 18.4 |
| No structured value in the final 24 h before **ICU** discharge, % (mechanism check) | 2.6 |
| Per-hospital % absent before hospital discharge, median (IQR) | 53.7 (31.6-67.3) |
| Median structured assessments per stay, No. | 55 |

Structured neurological assessments were charted densely within the intensive care unit (median 55 per stay; absent before ICU discharge in only 2.6%) but were absent in the final 24 hours before hospital discharge in 47.9% of stays (per-hospital median 53.7%). The end-of-stay gap observed in MIMIC-IV is therefore echoed across 187 hospitals as a coverage pattern: it localizes to the period after intensive care transfer rather than to undercharting within the intensive care unit, and is a property of ICU-database coverage rather than an independently validated hospital-wide documentation gap.

#### eTable 14. Timestamped-note companion analysis (MIMIC-III): temporal ordering of prognostic language

Acute brain injury admissions in MIMIC-III (11,079 admissions; 9,794 patients); prognostic language was detected in 1,288 admissions at the day level and 781 admissions among minute-resolution notes. All quantities are temporal orderings and co-occurrences of documentation, not causal effects. Because MIMIC-III discharge summaries carry only a date (no minute timestamp), Aim 1 uses a day-level ordering so that discharge summaries are eligible to be first.

| Measure | Value |
| --- | --- |
| **Aim 1: location and timing of the earliest prognostic-language statement (day-level)** |  |
| Earliest prognostic language in a timestamped clinical note (first or exclusively), % | 57.4 |
| Earliest prognostic language in the discharge summary only, % | 39.2 |
| Documented in a timestamped note (any time), % | 60.8 |
| Documented in the discharge summary (any time), % | 58.4 |
| Among admissions with both (n = 247), timestamped note precedes discharge summary, median (IQR), d | 2.0 (1.0-6.0) |
| Timestamped note strictly earlier than the discharge summary, % | 82.2 |
| **Aim 2: recency of command-following at the first prognostic statement** (n = 781 with prognostic language among minute notes) |  |
| Any command-following documented before the statement, % | 61.3 |
| Most recent such documentation positive, % (of those with prior documentation) | 54.3 |
| Command-following documented within 6 h before the statement, % | 17.5 |
| Command-following documented within 12 h before, % | 21.5 |
| Command-following documented within 24 h before, % | 27.9 |
| Command-following documented within 48 h before, % | 34.1 |
| Median age of the most recent positive command-following, h (IQR) | 13.2 (0-49) |
| **Aim 3: ordering vs active limitation and death (note-based; see caveat)** |  |
| Prognostic language preceded first active-limitation note, % | 73.2 |
| Prognostic language with no documented active-limitation note, % | 30.3 |
| Among in-hospital deaths, statement preceded death by, median (IQR), h | 43.5 (14-104) |
| **Documentation coverage among in-hospital deaths** (n = 1,831 with minute notes) |  |
| Deaths with any documented prognostic language, % (95% CI) | 29.1 (27.0-31.2) |
| **Extractor precision in MIMIC-III note types** (blinded physician adjudication, n = 160/register) |  |
| Prognostic language, % genuine (95% CI) | 98.8 (95.6-99.7) |
| Command-following, % genuine (95% CI) | 100 (97.7-100) |
| Active limitation, % genuine (95% CI) | 68.1 (60.6-74.8) |
| Physician–analyst agreement (Gwet AC1): prognostic language 0.99, command-following 1.00, active limitation 0.65 |  |

Aim 2 shows that the "most recent positive command-following" is frequently stale: although it was positive in 54.3% of admissions with prior documentation, that observation was a median of 13 hours old and fell within the prior 24 hours in only 27.9%. **Caveat for Aim 3:** the active-limitation note register was only 65% precise on analyst adjudication (37.5% hypothetical or planning language, e.g., "plan versus comfort measures"); the note-based ordering is therefore reported for context only, and a structured, adjudicated code-status comparator (code-status transitions, adjusted for severity and comorbidity) is the basis for inference. Extractor precision was assessed by blinded adjudication of a stratified sample of MIMIC-III timestamped notes (160 flagged excerpts per register): a board-certified physician, blinded to the extractor label and note type, judged whether each matched phrase was a genuine instance of its register. Prognostic language and command-following were highly precise in these note types (98.8% and 100%); the note-based active-limitation register was less so (68.1%, most errors being planning or hypothetical language), which is why the primary Aim-3 outcome uses structured code-status transitions rather than the note register. Physician judgments agreed closely with an independent analyst adjudication (overall Gwet AC1 0.91). Because prognostic language is documentation rather than the underlying clinical event and severity is unmeasured, these orderings describe the record and do not support causal inference.

#### eTable 15. Code-status limitation association in the timestamped cohort (MIMIC-III)

Exposure: first documented prognostic-language statement (physician-adjudicated register; eTable 14). Outcome: first structured code-status transition from full code to a limitation recorded in chartevents, as a composite and by component (comfort measures; do-not-resuscitate/do-not-intubate without comfort measures). Death and discharge are competing events. All hazard ratios are unpenalized cause-specific estimates with 95% CIs clustered by patient, adjusted for the expanded covariate set (age, primary phenotype, worst Glasgow Coma Scale motor score, mechanical ventilation, deepest Richmond Agitation-Sedation Scale value, van Walraven comorbidity index, metastatic cancer, dementia, chronic liver disease, chronic renal disease, emergency admission, pre-landmark intensive care unit hours, and database source). Landmark models restrict to admissions whose most recent observed code-status state at the landmark is full code.

| Analysis | At risk, No. (patients) | With language, No. (events) | Total events, No. | Adjusted HR (95% CI) | E-value |
| --- | --- | --- | --- | --- | --- |
| Any limitation, 24-h landmark (primary) | 5,493 (5,171) | 100 (42) | 574 | 4.3 (2.9-6.5) | 8.1 |
| Comfort-measures transition, 24 h | 5,512 (5,188) | 102 (18) | 247 | 3.9 (2.3-6.7) | 7.3 |
| DNR/DNI without comfort, 24 h | 5,493 (5,171) | 100 (32) | 458 | 4.2 (2.7-6.6) | 7.9 |
| Higher-risk neurologic subgroup, any, 24 h | 3,102 (NA) | 86 (39) | 504 | 4.4 (2.9-6.8) | 8.3 |
| Covariates measured pre-statement, 24 h | 5,493 (5,171) | 100 (NA) | 574 | 4.6 (3.3-6.3) | 8.6 |
| Adjusted for pre-landmark note volume, 24 h | 5,493 (5,171) | 100 (42) | 574 | 4.3 (2.9-6.5) | 8.1 |
| Adjusted for acute physiology, 24 h | 5,493 (5,171) | 100 (42) | 574 | 4.1 (2.7-6.2) | 7.6 |
| Acute physiology with missing-data indicators, 24 h | 5,493 (5,171) | 100 (42) | 574 | 3.6 (2.5-5.3) | 6.6 |
| Pseudo-index, equal covariate window | 4,375 (NA) | 429 (NA) | 448 | 6.2 (4.9-7.8) | 11.9 |
| Incidence-density risk-set match, clustered | 2,574 (NA) | 429 (NA) | 322 | 5.4 (3.9-7.3) | 10.2 |
| Incidence-density risk-set match, matched-set stratified | 2,574 (NA) | 429 (NA) | 322 | 3.2 (2.9-3.6) | 5.9 |
| Any limitation, 48-h landmark | 5,629 (5,280) | 117 (47) | 502 | 4.4 (3.0-6.4) | 8.2 |
| Any limitation, whole-stay time-varying | 7,573 | NA (1,137) | 1,137 | 3.8 (3.2-4.4) | 7.0 |
| Time-varying, 12-h statement-order interval | 7,573 | NA (1,137) | 1,137 | 3.3 (2.8-3.9) | 6.0 |
| First-admission only, 24 h | 5,013 | 92 (NA) | NA | 4.4 (3.2-6.2) | 8.3 |
| Discharge alive (competing-outcome contrast), 24 h | 5,493 | 100 (NA) | NA | 0.9 (0.6-1.3) | NA |

The crude 24-hour hazard ratio was 6.7; the sparser 5-covariate model gave 3.5, so the expanded adjustment did not attenuate the association but slightly increased it (to 4.3). The higher-risk neurologic subgroup (intracranial hemorrhage, subarachnoid hemorrhage, subdural hemorrhage, anoxic injury, cardiac arrest, or any mechanical ventilation) gave a similar estimate (any limitation 4.4; comfort measures 3.9), so the association is not an artifact of pooling milder admissions. When covariates for exposed admissions were measured strictly before the first prognostic statement (and before the landmark for unexposed admissions), the estimate was similar or slightly higher (4.6), arguing against reverse-time contamination of the adjustment set. Adjusting further for pre-landmark documentation volume (the total and physician note counts before the landmark) left the estimate unchanged (4.3), so the association is not an artifact of exposed admissions being more heavily documented. In an equal-covariate-window analysis that assigned each unexposed admission a pseudo-index time sampled from the exposed statement-time distribution and measured covariates for both groups only before their own index (4,375 admissions; 429 exposed; 448 events), the association was not attenuated (HR, 6.2; 95% CI, 4.9-7.8), indicating that the unequal pre-statement window did not generate the finding. Adjustment for acute physiology (worst pre-landmark creatinine, bilirubin, platelets, lactate, blood urea nitrogen, and arterial oxygen tension, and any vasopressor exposure) changed the estimate only modestly (4.1; 3.6 with missing-data indicators for the selectively measured labs), so the association was not materially attenuated by the available acute-physiology variables. In a corrected incidence-density risk-set matched analysis (sampling up to 5 controls that were alive, admitted, full-code, limitation-free, and not-yet-exposed at each statement hour; controls were allowed to become exposed later and were censored at their own first statement), the association remained elevated: HR 5.4 (95% CI, 3.9-7.3) clustered by patient and 3.2 (95% CI, 2.9-3.6) with matched-set stratification (429 sets; 2,145 controls; 322 events). This corrects an earlier version that had restricted controls to never-exposed admissions, which used post-index information. Proportional hazards did not hold for the exposure (Schoenfeld P = .02): a piecewise model split at 72 hours gave an early hazard ratio of 4.5 (95% CI, 3.0-6.5; 0-3 days) and a later hazard ratio of 1.7 (95% CI, 0.9-3.1; >3 days), so the single hazard ratio summarizes a time-varying effect concentrated in the first days and not statistically distinguishable from the null thereafter. The unadjusted 14-day competing-risks cumulative incidence was 40% versus 8.5% for any limitation and 16% versus 3.5% for comfort measures (Figure 2). Among admissions with both a first statement and an order, the statement preceded the order in 56.9% (median lead, 37.6 hours; ≥12 hours in 45.1%), and the required 12-hour interval retained the association. The discharge-alive contrast is reported as a descriptive competing-outcome comparison rather than a strong falsification test, because a genuinely poor prognosis would be expected to reduce discharge alive; the non-prognostic text registers (Table 2; eTable 20) provide a stronger specificity check. The E-value is the strength of association an unmeasured confounder would need with both exposure and outcome to explain the estimate away; because the clinician's global prognostic judgment could plausibly reach this magnitude and is captured only in part despite adjustment for acute physiology (a full APACHE score was not computable), the E-value does not exclude residual confounding, and causal inference is not supported. NA denotes a quantity not defined for a given specification.

#### eTable 16. Characteristics of admissions included in versus excluded from the landmark risk set

The 24-hour landmark analysis restricts to admissions still alive, admitted, and limitation-free at 24 hours whose most recent observed code-status entry by 24 hours is full code (included). Admissions that met the survival and limitation-free conditions but had no observed full-code entry by 24 hours were excluded from the landmark risk set. This table characterizes that selection.

| Characteristic | Included (observed full code), n = 5,493 | Excluded (no observed full code), n = 1,584 |
| --- | --- | --- |
| Age, median, y | 62.4 | 66.0 |
| Prognostic language by 24 h, % | 7.4 | 8.0 |
| In-hospital death, % | 12.7 | 16.8 |
| Most frequent phenotype | Traumatic brain injury (29.6%) | Ischemic stroke (29.9%) |
| Second phenotype | Ischemic stroke (22.4%) | Status epilepticus (15.2%) |
| Third phenotype | Intracranial hemorrhage (14.0%) | Anoxic injury (14.6%) |

The included cohort was modestly younger and lower-mortality than the excluded cohort (12.7% vs 16.8% in-hospital death), consistent with excluded admissions being sicker or more often admitted already under a limitation, whereas exposure prevalence was similar in the two groups (7.4% vs 8.0%). The requirement of an observed full-code state therefore selects a somewhat healthier risk set but does not appear to select on the exposure itself.

#### eTable 17. Characteristics of the MIMIC-IV documentation-mapping cohort (admissions with a discharge summary), overall and by presence of prognostic language

Data are No. (%) unless otherwise noted. This is the cross-sectional cohort for the documentation-mapping analyses (localization, responsiveness documentation, and co-occurrence); the code-status association is analyzed in the timestamped MIMIC-III landmark cohort (main Table 1). Race and ethnicity distributions and adjusted associations are reported in eTable 9.

| Characteristic | Overall | Prognostic language present | Prognostic language absent |
| --- | --- | --- | --- |
| No. | 23,239 | 1,127 | 22,112 |
| Age, median (IQR), y | 69 (57-81) | 71 (59-82) | 69 (56-81) |
| Male | 12,650 (54.4) | 600 (53.2) | 12,050 (54.5) |
| ICU admission | 12,058 (51.9) | 964 (85.5) | 11,094 (50.2) |
| In-hospital death | 2,864 (12.3) | 816 (72.4) | 2,048 (9.3) |
| Discharged home | 9,564 (41.2) | 43 (3.8) | 9,521 (43.1) |
| To rehabilitation | 3,725 (16.0) | 32 (2.8) | 3,693 (16.7) |
| To hospice | 463 (2.0) | 81 (7.2) | 382 (1.7) |

Phenotype distribution (No., %): cardiac arrest 1,846 (7.9); anoxic-ischemic injury 2,922 (12.6); intracerebral hemorrhage 2,242 (9.6); subarachnoid hemorrhage 935 (4.0); subdural hemorrhage 916 (3.9); acute ischemic stroke 7,835 (33.7); traumatic brain injury 5,878 (25.3); status epilepticus 252 (1.1); central nervous system infection 413 (1.8).

#### eTable 18. Covariate completeness in the 24-hour landmark cohort, by prior prognostic language

Charting completeness of the adjustment covariates before the 24-hour landmark. When a value was not charted before the landmark, the covariate was set to the cohort median; no admission was excluded for missing covariates. Mechanical ventilation and vasopressor exposure are observed indicators (absence denotes not ventilated / no vasopressor, not missing).

| Covariate | Charted before landmark, exposed (n = 100), % | Charted before landmark, unexposed (n = 5,393), % |
| --- | --- | --- |
| GCS motor score | 100.0 | 99.9 |
| Richmond Agitation-Sedation Scale | 4.0 | 19.3 |
| Serum creatinine | 99.0 | 99.8 |
| Blood urea nitrogen | 99.0 | 99.8 |
| Platelet count | 99.0 | 99.7 |
| Arterial oxygen tension | 93.0 | 61.0 |
| Serum lactate | 81.0 | 62.7 |
| Total bilirubin | 63.0 | 47.8 |
| Mechanically ventilated (observed) | 86.0 | 49.7 |
| Received vasopressor (observed) | 36.0 | 17.7 |

The Glasgow Coma Scale motor score, creatinine, blood urea nitrogen, and platelet count were nearly always charted; the Richmond Agitation-Sedation Scale was sparse (charted in 19.3% of unexposed and 4.0% of exposed admissions) and was therefore frequently imputed. Lactate, arterial oxygen tension, and bilirubin were measured more often in the exposed (sicker) group, consistent with selective measurement in the most severely ill, so median imputation for these labs could understate their adjustment value. A sensitivity model that added missing-data indicators for lactate, arterial oxygen tension, and bilirubin gave a modestly lower estimate (HR, 3.6; 95% CI, 2.5-5.3; eTable 15), still elevated. The sedation-charting sparsity and selective laboratory measurement are stated as limitations.

#### eTable 19. Code-status association stratified by documentation era and phenotype (MIMIC-III, 24-hour landmark)

The primary physiology-adjusted 24-hour landmark model, fitted within strata. A small ridge penalty (0.1) was used to stabilize the sparser strata. Confidence intervals are clustered by patient.

| Stratum | At risk, No. | With language, No. | Events, No. | Adjusted HR (95% CI) |
| --- | --- | --- | --- | --- |
| CareVue era | 3,658 | 69 | 457 | 3.7 (2.5-5.5) |
| MetaVision era | 1,835 | 31 | 117 | 2.7 (1.4-5.3) |
| Intracranial hemorrhage | 767 | 32 | 104 | 3.2 (1.7-5.9) |
| Traumatic brain injury | 1,627 | 19 | 129 | 3.9 (1.8-8.6) |
| Anoxic injury or cardiac arrest | 472 | 21 | 111 | 3.1 (1.7-5.8) |

The association was present in both documentation eras and in each major phenotype, so it does not depend on a single charting system or injury type.

#### eTable 20. Specificity of the association: non-prognostic text registers (MIMIC-III, 24-hour landmark)

Full results for Table 2. Each row is the identical adjusted 24-hour landmark cause-specific Cox model with the exposure replaced by the first-occurrence of the register; confidence intervals clustered by patient; death and discharge competing.

| Text register (exposure) | Documented by 24 h, % | With register, No. | Events, No. | Adjusted HR (95% CI) | E-value |
| --- | --- | --- | --- | --- | --- |
| Prognostic language | 1.8 | 100 | 574 | 4.3 (2.8-6.4) | 8.0 |
| Radiographic severity | 83.0 | 4,560 | 574 | 1.1 (0.9-1.4) | 1.4 |
| Diagnostic-certainty hedging | 85.4 | 4,689 | 574 | 1.1 (0.9-1.4) | 1.4 |
| Generic critical-illness acuity | 16.8 | 922 | 574 | 1.3 (1.1-1.7) | 1.9 |

Despite being documented in 17% to 85% of admissions (versus 1.8% for prognostic language), radiographic-severity and diagnostic-hedging language were not associated with a subsequent code-status limitation and generic critical-illness language was only weakly associated (HR 1.3). This supports relative specificity of the prognostic-language association rather than an artifact of severity description, interpretive hedging, or documentation density; the registers nonetheless differ in prevalence, timing, and semantics, so the comparison is supportive, not definitive.

#### eTable 21. Two-physician adjudication of early statement-to-order sequences (MIMIC-III)

Two physicians independently classified the 143 admissions whose first structured code-status limitation occurred within 48 hours after the first documented prognostic-language statement, blinded to each other and to the automated first pass, into one category describing the relationship between the statement and the order.

| Category | Rater 1, No. (%) | Rater 2, No. (%) |
| --- | --- | --- |
| Statement preceded a separate later decision | 75 (52.4) | 54 (37.8) |
| Same goals-of-care episode | 41 (28.7) | 53 (37.1) |
| Prior directive or family preference | 11 (7.7) | 7 (4.9) |
| Clinical deterioration between statement and order | 6 (4.2) | 20 (14.0) |
| Timing or relationship unclear | 6 (4.2) | 2 (1.4) |
| Retrospective documentation | 4 (2.8) | 3 (2.1) |
| Copied-forward language | 0 (0) | 4 (2.8) |

The physicians agreed on the sequence classification in 60.8% of cases (87 of 143; Cohen κ, 0.43, moderate). Among those agreements, 52% (45 of 87) were classified as a prognostic statement preceding a separate later decision and 37% (32 of 87) as co-documentation of a single goals-of-care episode. The moderate agreement is itself informative: even on manual chart review the temporal pairing is a genuine sequence in some admissions and co-documentation of one conversation in others, so even manual review could not reliably distinguish influence from co-documentation. This adjudication concerns the statement-to-order relationship and is distinct from the extractor-precision adjudication in eTable 4.

### eFigures

#### eFigure 1. Prognostic language: prevalence and note-stream localization (MIMIC-IV)

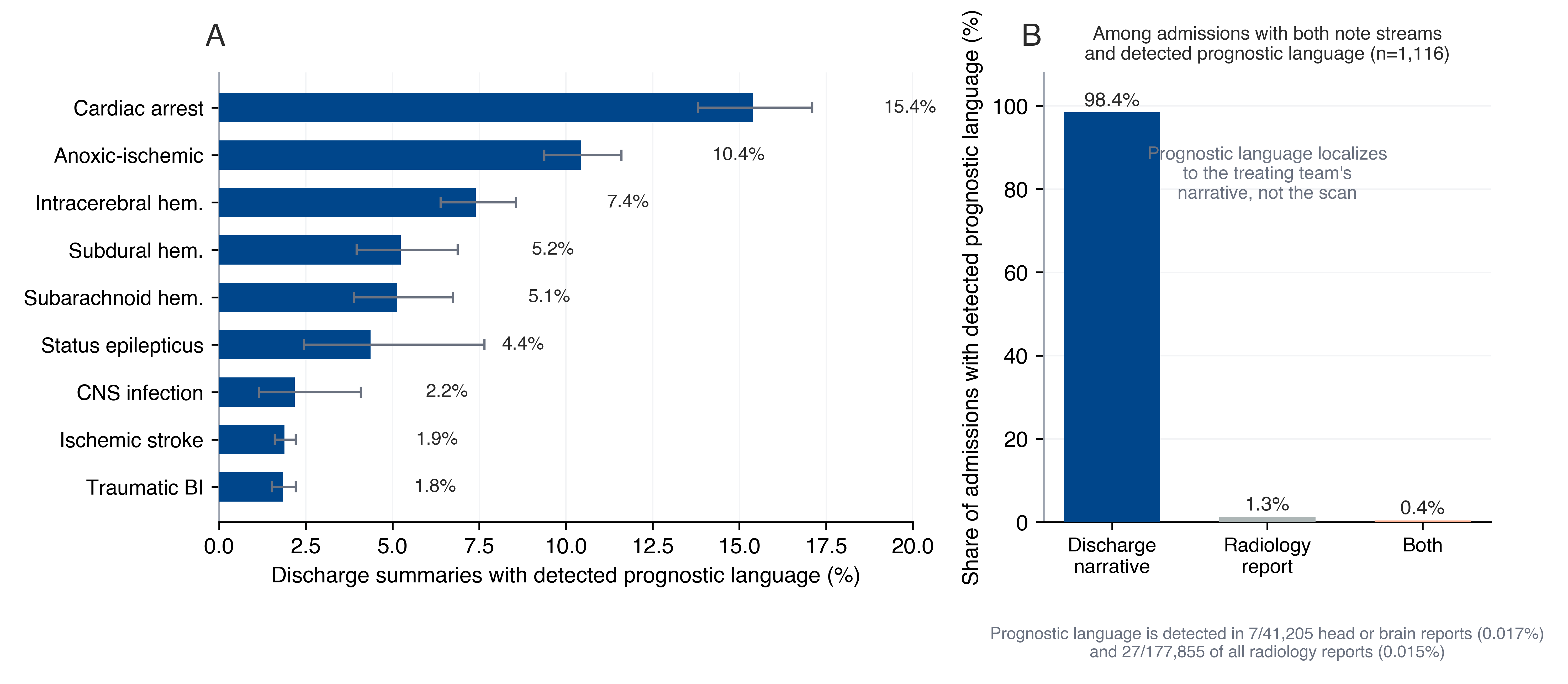

(A) Prevalence of detected prognostic language in discharge summaries by phenotype (Wilson 95% CIs). (B) Localization of detected prognostic language across note streams among admissions with both streams available and prognostic language present (n = 1,116); across all radiology reports in the cohort, the extractor detected prognostic language in 27 of 177,855 (0.015%).

#### eFigure 2. Documentation of responsiveness (MIMIC-IV)

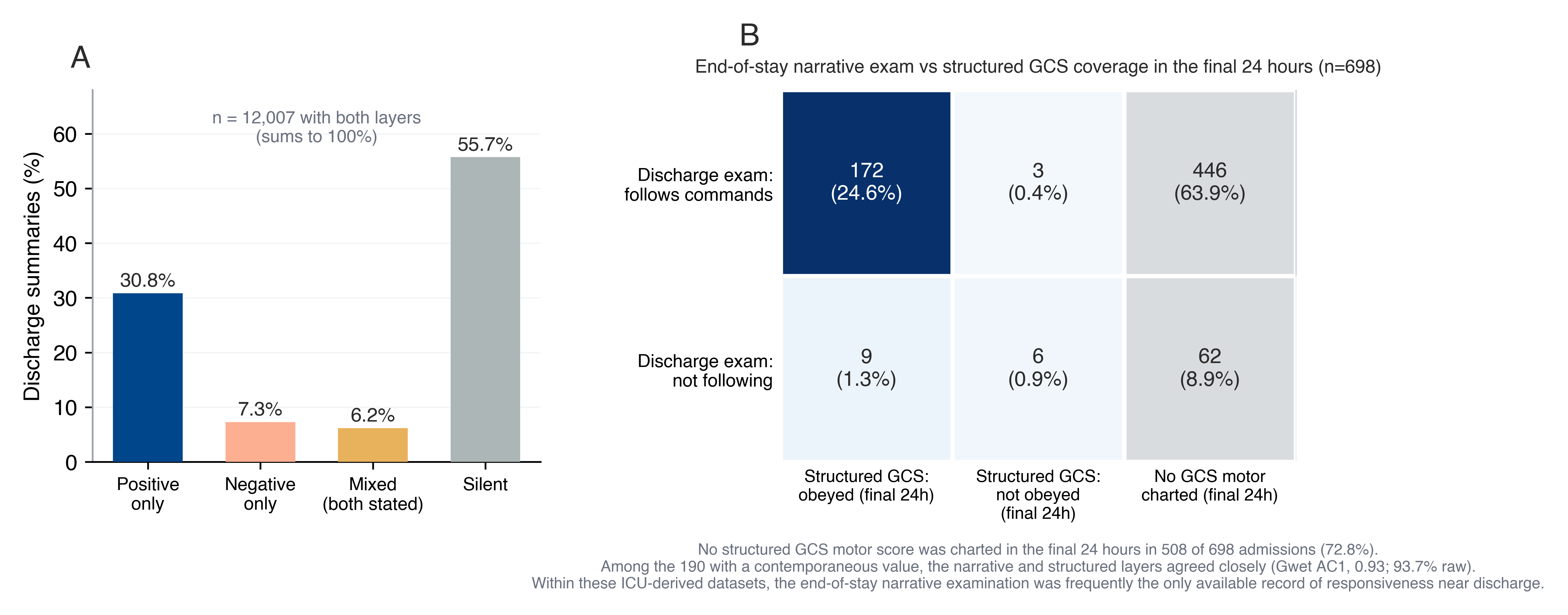

(A) Mutually exclusive distribution of narrative command-following documentation among admissions with both layers (n = 12,007): positive only, negative only, mixed, and silent. (B) Coverage of structured Glasgow Coma Scale motor scores in the final 24 hours among end-of-stay-examined admissions (n = 698), cross-classified by narrative examination (follows commands vs not following) and structured state (obeyed, documented not-obeying, or no GCS motor charted). No final-24-hour GCS motor score was charted in 508 admissions (72.8%); among the 190 with a contemporaneous value, agreement was high (Gwet AC1, 0.93; 93.7% raw).
